# Lower Cancer Incidence Three Years After COVID-19 Infection in a Large Veteran Population

**DOI:** 10.1101/2025.01.14.25320527

**Authors:** Jerry Bradley, Fei Tang, Natasha Resendes, Dominique Tosi, Iriana Hammel

## Abstract

**Background:** The role of COVID-19 infection in cancer incidence risk is not known. COVID-19 infection could lead to increased cancer risk, as seen with other viruses, or it could lead to decreased risk due to the activation of the immune response during acute infection. This study aimed to determine the association between cancer incidence in US Veterans after COVID-19 infection.

**Methods:** We conducted a retrospective cohort study of US Veterans comparing those who tested positive for COVID-19 during the first wave of COVID-19 between March 15, 2020, and Nov 30, 2020, to those who tested negative. We used data from the COVID-19 Shared Data Resource and used Cox proportional hazard regression models to determine the hazard ratio of a new cancer diagnosis within a three-year follow-up period for the COVID-19 positive patients compared to those who were negative. Covariates included age, race, ethnicity, sex, BMI, smoking, being an active patient in the VHA system in the past 12 months of the COVID-19 test, and other factors.

**Results:** 516156 patients were included in this study, with 88590 (17.2%) COVID-19 positive, 427566 (82.8%) COVID-19 negative. The ages of the COVID-19 positive and negative patients were 57.8±16.4 and 59.4±15.8, respectively. For those who survived for at least 30 days after COVID-19 testing, COVID-19 infection was associated with a 32% reduction in the hazard of cancer. The reduction of the hazard was similar across sexes and races, except in Asians. Above 45 years of age, the hazard of cancer incidence further decreases with advancing age.

**Conclusions:** Patients who were diagnosed with COVID-19 in the first wave of the pandemic had a decreased risk of cancer incidence in a 3-year follow-up across gender and race. Further multicenter prospective cohort studies are needed to evaluate the mechanism of this interaction.

## Introduction

The COVID-19 pandemic has introduced a new viral factor with relatively unknown effects on cancer development. Approximately 10% of cancers worldwide are virus-induced, with Epstein-Barr virus (EBV), Hepatitis C virus (HCV), and human papillomavirus (HPV) being examples of known viral oncogenic drivers (1). The mechanism by which viruses lead to cancer development might be driven by the dysregulation of signaling pathways and alterations in host DNA and RNA processes (2-4). It is unknown if, during the acute infection state of COVID-19, dysregulation in immune signaling (5, 6), DNA damage with activation of pro-inflammatory processes (7), or other mechanisms might increase the risk of cancer development.

Early in the pandemic, researchers were aware that COVID-19 induced morphological and inflammatory changes in peripheral blood monocytes (8). However, the significance of these changes has been debated. A recent analysis by Dr. Bharat et al. found that COVID-19 could induce non-classical monocytes to act with anticancer properties (9). Specifically, they found that COVID-19 single-stranded RNA (ssRNA), unlike influenza ssRNA, could increase the population of this monocyte line. (9). The implication of these changes as a function of monocyte exposure to COVID-19 ssRNA raises the question of whether contracting COVID-19 may affect cancer development in the short term.

Our team set out to compare the incidence of cancer in patients with COVID-19 to those without. We hypothesized that patients would have a reduced risk of cancer across the board in patients who were exposed to COVID-19 as compared to non-exposed patients.

## Methods

### Study Design, Data Sources, and Study Population

We conducted a retrospective cohort study to assess the association between COVID-19 infection and new cancer diagnoses in Veterans who were free of cancer at baseline. We included Veterans who tested positive for the first time for SARS-CoV-2 (polymerase chain reaction or antigen test) in VA medical centers or clinics between March 15, 2020, and November 30, 2020 (Alpha/Beta waves). The control group included Veterans who had a negative test result in this time frame and did not report positive test results in the next three years. We excluded patients who had any cancer diagnosis at baseline and those who died within 30 days after SARS-CoV-2 infection. We used nationwide data from the VHA medical centers from the VA COVID-19 Shared Data Resource. The Miami Veterans Affairs Healthcare System Institutional Review Board approved this study. The IRB granted a waiver for informed consent. The data was fully anonymized according to VHA standards prior to analysis. The data was accessed on 01/10/2024 to 17/12/2024.

### Study Outcome

The primary outcome was any cancer diagnosis after COVID-19 testing (positive or negative) as documented by ICD-10 codes.

### Covariates

Age, race, ethnicity, sex, BMI, smoking, kidney disease, COPD, diabetes, hypertension, hyperlipidemia, obesity, chronic lung disease, PTSD, alcohol dependence, drug dependence, being an active patient in the past 12 months, exposure to toxic substances, other viral infections such as a history of exposure to Hepatitis B, Hepatitis C, cytomegalovirus, mononucleosis, HIV among others, and immunodeficiency conditions secondary to prior history transplant or conditions related to immune response.

### Statistical Analysis

Continuous variables are presented as mean ± standard deviation, median with interquartile range, and categorical variables as frequencies and percentages. When information on race, ethnicity and smoking status was not available, we reported the data as “Unknown.” The Cox proportional hazard model was used to evaluate the association between COVID-19 and cancer diagnosis, adjusting for the covariates listed above. The proportional hazards assumption was assessed and verified by testing the correlation between the Schoenfeld residuals and survival time. Time-to-event analyses began 30 days after the date of infection, and patients were censored on the date of death or at the end of the years of follow-up. The association was also assessed in patients of different age groups, sexes, and races. Statistical analysis was performed using R (R project for statistical computing, version 4·0·5).

## Results

### Baseline Characteristics

A total of 516,156 Veterans with SARS-CoV-2 testing in 2020 were included in this retrospective cohort study, with 346,084 (67.1%) white, 112,476 (21.8%) black, 44,392 (8.6%) Hispanic, 45,5388 (88.2%) males. A total of 88,590 (17.2%) patients tested positive for SARS-CoV-2. The baseline characteristics of the study cohort are summarized in Table 1. The mean age of the entire cohort was 59.1±16.0 years (median 62 years), with COVID-19 positive and negative patients having similar ages (57.8±16.4 vs 59.4±15.8). The COVID-19 positive and negative patients were similar in terms of other baseline characteristics, as well.

**Table 1.** Baseline Characteristics.

|  | **Total**<br>**n = 516156 (100%)** | **COVID-19 Negative**<br>**n = 427566 (82.8%)** | **COVID-19 Positive**<br>**n = 88590 (17.2%)** |
| --- | --- | --- | --- |
| **Age, mean (years) ±SD**<br>**(median; IQR)** | 59.1±16.0 (62; 48-71) | 59.4±15.8 (62; 49-71) | 57.8±16.4 (59; 45-71) |
| **Age Groups, n (%)** |  |  |  |
| **19-64** | 220523 (42.7%) | 185873 (43.5%) | 34650 (39.1%) |
| **$\geq 65$** | 295633 (57.3%) | 241693 (56.5%) | 53940 (60.9%) |
| **Male sex, n (%)** | 455388 (88.2%) | 376833 (88.1%) | 78555 (88.7%) |
| **Race, n (%)** |  |  |  |
| **White** | 346084 (67.1%) | 288910 (67.6%) | 57174 (64.5%) |
| **Black** | 112476 (21.8%) | 91545 (21.4%) | 20931 (23.6%) |
| **Asian** | 6256 (1.2%) | 5397 (1.3%) | 859 (1.0%) |
| **American Indian or**<br>**Alaska Natives** | 4378 (0.8%) | 3496 (0.8%) | 882 (1.0%) |
| **Native Hawaiian or**<br>**Other Pacific Islander** | 4783 (0.9%) | 3919 (0.9%) | 864 (1.0%) |
| **Unknown** | 42179 (8.2%) | 34299 (8.0%) | 7880 (8.9%) |
| **Ethnicity, n (%)** |  |  |  |
| **Hispanic** | 44392 (8.6%) | 34365 (8.0%) | 10027 (11.3%) |
| **Not Hispanic** | 457677 (85.7%) | 381616 (89.3%) | 76061 (85.9%) |
| **Unknown** | 14087 (2.7%) | 11585 (2.7%) | 2502 (2.8%) |
| **Smoking, n (%)** |  |  |  |
| **Current** | 109559 (21.2%) | 98650 (23.1%) | 10909 (12.3%) |
| **Former Smoker** | 192443 (37.3%) | 157933 (36.9%) | 34510 (39.0%) |
| **Never** | 169205 (32.8%) | 134629 (31.5%) | 34576 (39.0%) |
| **Unknown** | 44949 (8.7%) | 36354 (8.5%) | 8595 (9.7%) |
| **Comorbidity, n (%)** |  |  |  |
| **Kidney Disease** | 101248 (19.6%) | 84599 (19.8%) | 16649 (18.8%) |
| **COPD** | 84592 (16.4%) | 73056 (17.1%) | 11536 (13.0%) |
| **Diabetes** | 153007 (29.6%) | 125141 (29.3%) | 27866 (31.5%) |
| **Hypertension** | 296271 (57.4%) | 246537 (54.5%) | 49734 (54.8%) |
| **Chronic Lung Disease** | 160078 (31.0%) | 135996 (31.8%) | 24082 (27.2%) |
| **Overweight** | 35361 (6.9%) | 27676 (8.7%) | 7685 (6.5%) |
| **Toxic exposure** | 3911 (0.8%) | 3321 (0.8%) | 590 (0.7%) |
| **Alcohol Dependency** | 123841 (24.0) | 104676 (24.5%) | 19165 (21.6%) |
| **Drug Dependency** | 35969 (7.0%) | 32203 (7.5%) | 3766 (4.3%) |
| **Immunodeficiency** | 14572 (2.8%) | 12442 (2.9%) | 2130 (2.4%) |
| **Mortality** | 88414 (17.1%) | 72621 (16.4%) | 15793 (16.9%) |

### Association of COVID-19 Infection and Decreased Cancer Risk

Having a positive COVID-19 infection was associated with a 25% decrease in the hazard of having a cancer diagnosis in the three years follow-up (HR=0.75, 95% CI: 0.74-0.77), controlling for age, race, ethnicity, sex, BMI, smoking, kidney disease, COPD, diabetes, hypertension, hyperlipidemia, obesity, chronic lung disease, PTSD, alcohol dependence, drug dependence, being an active patient in the past 12 months, exposure to toxic substances, other viral infections, and immunodeficiency.

### Association of COVID-19 Infection and Decreased Cancer Risk by Age Groups, Gender, and Race

We assessed the association between positive COVID-19 infection and the risk of having a cancer diagnosis within three years in different age groups. We found that the older groups showed a greater reduction in the hazard of new cancer diagnosis. For the age group of 85-99, having a positive COVID-19 infection was associated with a 36% decrease in the hazard of having a cancer diagnosis in the three years follow-up (aHR: 0.64, 95% CI: 0.58-0.70), for age group of 75-84, the reduction was 26% (aHR: 0.74, 95% CI: 0.70-0.78); and for the age group of 55-64, the reduction was 16% (aHR: 0.84, 95% CI:0.80-0.89) (Table 2). Regarding sex, the reduction in new cancer diagnosis was slightly higher in men (aHR: 0.74, 95% CI: 0.70-0.78) than in women (aHR: 0.85, 95% CI: 0.76-0.94) (Table 3). Regarding race and ethnicity, the reduction in new cancer diagnoses was similar between white, black, Asian, and Hispanic populations (Table 4).

**Table 2.** aHR for New Cancer Diagnosis within three years for Patients with Positive COVID-19 Testing Compared to Patients of Negative Testing by Age.

|  | **Number of events (%)** | **aHR (95% CI)** |
| --- | --- | --- |
| 18-24 (n=3549) | 17 (0.5%) | 1.16 (0.37-3.63) |
| 24-34 (n=44690) | 383 (0.9%) | 0.85 (0.65-1.12) |
| 35-44 (n=61155) | 1130 (1.8%) | 0.93 (0.80-1.09) |
| 45-54 (n=73400) | 3334 (4.5%) | 0.87 (0.79-0.95) |
| 55-64 (n=112828) | 11055 (9.8%) | 0.84 (0.80-0.89) |
| 65-74 (n=146407) | 25956 (17.7%) | 0.76 (0.73-0.79) |
| 75-84 (n=53915) | 11549 (21.4%) | 0.74 (0.70-0.78) |
| 85-99 (n=20201) | 4155 (20.6%) | 0.64 (0.58-0.70) |

**Table 3.** aHR for New Cancer Diagnosis within three years for patients with positive COVID-19 testing compared to patients with negative testing by sex.

|  | aHR (95%CI) |
| --- | --- |
| Man | 0.74 (0.73-0.76) |
| Women | 0.85 (0.76-0.94) |

**Table 4.**
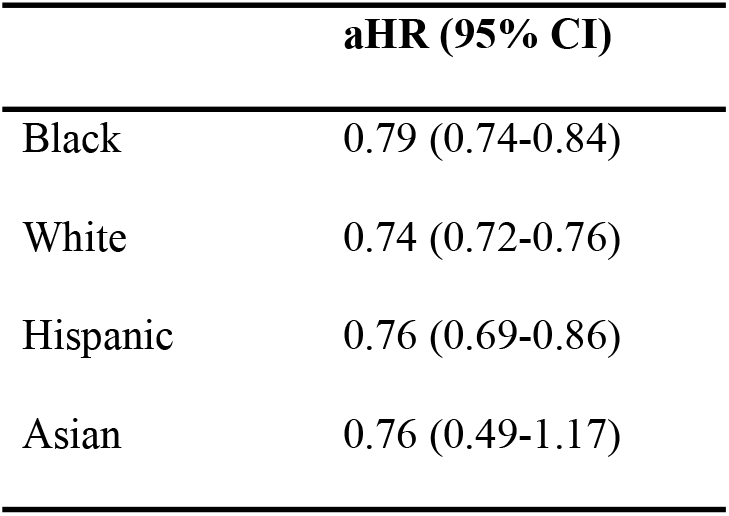
aHR for New Cancer Diagnosis within three years for Patients with Positive COVID-19 Testing Compared to Patients of Negative Testing by Race and Ethnicity.

## Discussion

### COVID-19 Exposure and Cancer Risk

Our findings show that beyond the age of 45, exposure to COVID-19 is associated with a reduction in cancer incidence across all major cancer types. It is plausible that the immunologic changes that occur during acute COVID-19 infection (10, 11) might lead to the activation of monocytes with anticancer properties, as has been reported by other investigators (9), or another process may lead to heightened immunity that has not been identified. The inflammatory response to COVID-19 may have a short-term protective benefit that wanes over time. Therefore, it is uncertain how long this risk reduction might be observed in the long-term beyond the 3-year period of this study. Further research and longer follow-up periods are required to fully understand the mechanisms responsible for the reduction in cancer incidence.

### Pandemic Healthcare Disruptions

COVID-19 has introduced several new healthcare disruptions in cancer surveillance. Cancer trends during the pandemic showed an initial reduction, followed by a general return to the pre-pandemic levels (12, 13). These changes were expected, given the drastic reduction in preventive services in the US (14). An analysis of trends in US cancer rates revealed the highest variability in breast, cervical, prostate, and colorectal cancers, likely driven by a reduction in surveillance screening (15). For this study, patients were analyzed for three years after the initial index infection. This timeframe was extended to include the post-pandemic return to standard screening; hence, the reduction we saw in our results is likely not a reflection of reduced screening or missed diagnosis.

### Age and Racial Differences

Racial and sex disparities are known to exist after COVID-19 infection, with men being more likely to have adverse outcomes after infection (16). These racial and gender disparities have improved over time (17) but remain an active area of investigation. Our findings showed that the overall rate of cancer reduction was the same regardless of race or sex, except for Asians, who comprised a small percentage of VA users. These findings were unexpected given the known racial and ethnic disparities in cancer incidence (18). The relative uniformity of this reduction across both sexes and races suggests the potential of a biological process independent of these factors.

Regarding age, the incidence of cancer appeared to decrease with each decade of life in the COVID-19 group compared to that in the non-exposed group. This is surprising, given that cancer diagnoses typically increase with age. Several factors may contribute to this finding. Older patients have a higher rate of death than younger patients with COVID-19 (19). Our study examined patients during the initial alpha and delta waves. During the pandemic, mortality rates reached their highest point in the delta-wave cohort (20). We attempted to mitigate the risk of survival bias by censoring and multivariate analyses for potential confounders. Our study showed that mortality rates were similar between groups after the 30-day period. However, several factors remain that cannot be adjusted through statistical processes. Given the reduction in cancer rates across the board in the infected cohort, patients in this group who survived the initial infection may have been at a lower risk for cancer due to environmental or genetic reasons. Similarly, the increased mortality in this group could have conferred a survival advantage regarding cancer risk. Therefore, care must be taken when attempting to link viral exposure to reduced cancer outcomes, as these findings do not support causation.

### Strengths and Limitations

Given the retrospective and observational nature of this study, the findings should be interpreted with caution. One of the strengths of this study is that it comprises a large nationwide database in which multiple risk factors and outcomes can be analyzed. An initial limitation of the study is that this is only a 3-year cohort analysis of cancer incidence in a Veteran population that is mostly comprised of male patients. These findings might change as five and ten-year data become available and may be different in non-Veteran populations. Additionally, we attempted to count for re-infection data; however, we were unable to capture all cases due to the increased availability of COVID-19 testing outside of the VA (at-home testing, state, and county testing sites, among others) during subsequent re-infections and potentially milder disease or other medical and socioeconomic reasons for which veterans did not seek medical attention. It is unknown how reinfection alters the risk of cancer development in these patients, and the extent to which it might induce other physiological changes.

## Conclusion

Despite the limitations of our retrospective observational study, cancer incidence appears to be lower in patients diagnosed with COVID-19 across all major cancers and independent of race or sex. Further research is required to determine the precise causes and mechanisms underlying these findings.

## Data Availability

The datasets presented in this article are not readily available because of the Department of Veterans Affairs data policies. Requests to access the datasets should be directed to the Department of Veterans Affairs

## Acknowledgments

We would like to acknowledge the support of the Miami VAMC, Geriatric Research Education and Clinical Center, and the US Veterans who served as our study population. This study was supported using resources and facilities of VINCI, VA HSR RES 13-457

